# Aggregation of mortality data by place and cause mask underlying trends in COVID-19 excess mortality

**DOI:** 10.64898/2026.09.04.26362260

**Authors:** Kelly Herdzik, Krzysztof Sakrejda, Abram Wagner, Jon Zelner

**Affiliations:** Department of Global Health, Amsterdam University Medical Center, Amsterdam, Netherlands; Department of Epidemiology, University of Michigan School of Public Health, Ann Arbor, MI USA; Center for Social Epidemiology & Population Health, University of Michigan School of Public Health, Ann Arbor, MI USA

## Abstract

Difficulties ascertaining the true burden of infection and mortality from SARS-CoV-2 hindered disease surveillance and healthcare resource allocation throughout the COVID-19 pandemic.

Although all-cause excess mortality estimates have long been deployed to avoid the challenges of case detection when testing availability is variable, aggregation of mortality by cause of death and spatial unit may mask underlying patterns and hide underlying associations with social factors. Furthermore, some non-infectious causes of death may have decreased during the pandemic, resulting in all-cause excess mortality capturing a net pandemic effect rather than the direct burden of Covid-19.

Using mortality data in the state of Michigan from January 2015 – November 2019, we calculated the expected mortality for December 2019 – November 2022 using negative binomial regression. Comparing these estimates to the observed mortality, we calculated the burden of excess mortality during the winter 2019 influenza season and COVID-19 pandemic, as divided into 7 distinct periods based on dominant viral strain and pharmaceutical and non-pharmaceutical interventions implemented. All analyses were performed with the data aggregated by cause of death and to the county level and repeated at both the county and census tract level disaggregated into two crude cause of death categories: acute respiratory infections (ARIs) and all other causes (non-acute respiratory infections, non-ARIs). We examined spatial heterogeneity of excess mortality by cause using the Theil index to determine if variation in excess mortality was driven by differences between or within counties.

The excess mortality rate from ARI peaked in the first phase of the pandemic, with a ratio of 10.7 observed to expected deaths (95% CrI: 10.3 – 11.1). However, the all- cause excess mortality rate rose only to 1.20 excess deaths per expected (95% CrI: 1.17 – 1.22) and the non-ARI excess mortality rate was unchanged. Comparing ARI excess mortality rates calculated at the county vs. census tract level demonstrated large and unpredictable variation, with county level estimates ranging from 0.42 to 1.2 times the tract level estimates. However, county level estimates of non-ARI and all- cause excess mortality ratios were consistently closer to tract level estimates. Within county variation of ARI mortality decreased during the pandemic and slowly returned to reference levels, suggesting spatial patterning of ARI mortality became more similar across census tracts as rates rose statewide. Social vulnerability was associated with an increased ARI excess mortality rate ratio at the census tract level during the first pandemic phase (EMRR: 1.1, 95% CrI: 1.08 - 1.13), but this effect was attenuated when aggregated to all-cause mortality or to the county level.

Our findings indicate that using county-level all-cause excess mortality as a proxy for ARI excess mortality obscured the burden of ARI death, particularly during the most acute phases of the COVID-19 pandemic. Relying on aggregated metric of all-cause mortality limits the power to detect large shifts in cause-specific mortality, and that limitation can be mitigated by even using coarse categories such as ARI vs. non-ARI deaths. Similarly, finer scale spatial units allow for the detection of local trends necessary to identify associated social factors.

## Introduction

Difficulties in ascertaining the true burden of infection and mortality from SARS-CoV-2 have hindered disease surveillance and healthcare resource allocation efforts since the outset of the COVID-19 pandemic. In the face of continued transmission of SARS-CoV-2 and emergence of new acute respiratory infection (ARI) threats, such as H5N1 influenza, passive case reporting has proven insufficient to detect shifting epidemic dynamics. However, active testing and case-finding efforts are expensive, difficult to set up rapidly, and still subject to reporting biases.

Estimates of all-cause excess mortality have long been deployed to sidestep these challenges and provide a fuller picture of mortality in the aftermath of natural disasters such as earthquakes, hurricanes, and heatwaves, as well as during aberrant infectious disease events such as the 2009 H1N1 influenza pandemic (1–6).

Relative to infections, mortality is a well-defined event, and mortality due to infectious disease is typically medically supervised with more scrutiny than other medical events. There are also legal and cultural expectations for recording the cause of death, along with contributing causes. Because of this higher level of scrutiny, excess mortality analyses have been promoted as a ‘gold standard’ for assessing the total burden of death attributable to COVID-19 (7). However, the COVID-19 pandemic presents a unique set of circumstances: In addition to exacting a very high death toll, it is characterized by having a very long duration as well as socially and geographically heterogeneous impacts that are of primary interest for evaluation and intervention. In this analysis, we use data from the U.S. state of Michigan to show how these factors can make all-cause excess mortality an unreliable and noisy signal of temporal change and spatial heterogeneity in pandemic-associated mortality. We demonstrate that an alternative approach in which excess mortality is disaggregated into acute respiratory infection (ARI) and non-ARI deaths provides a clearer and more useful picture of temporal shifts and spatial heterogeneity in pandemic-related mortality than all-cause mortality alone.

## What does pandemic-related all-cause excess mortality tell us?

All-cause excess mortality ostensibly reflects the total effect, i.e. the combined direct and indirect effects, of an exposure on the burden of mortality in a population. This includes deaths stemming from the disaster or event of interest, as well as ‘collateral’ deaths associated with secondary impacts of the event. Examples of indirect deaths include those resulting from the emergence of infectious diseases like Cholera and Polio following the breakdown of infrastructure in the aftermath of a devastating earthquake or in a conflict zone (8,9). These may also reflect deaths from chronic and acute illnesses resulting from the inability to access primary care during periods of widespread disruption. The lack of specificity inherent in all-cause excess mortality estimates is often considered an asset, as it accounts for the entire set of deaths that would not have occurred in the counterfactual scenario where the event of interest had not happened.

The direct and indirect effects of a sustained, geographically diffuse exposure, like a global pandemic, that plays out over many months or even years challenge the utility of all-cause mortality because the balance of direct, infection-associated deaths and indirect deaths may change over time. More problematically, the directionality of these changes may differ, with deaths due to non-infection causes (e.g. automobile and workplace accidents) declining when infection-related mortality rates are highest. Combining these into a single all-cause estimate can obscure these effects and make evaluation of change over time and across geographic locations difficult.

Despite these issues, all-cause excess mortality has illuminated the magnitude of SARS-CoV-2 mortality at the national (10) and state levels (11) and provided granular insights into county-level impacts of the pandemic (12). While these studies have documented the shifting burden of aggregate mortality, we still know little about the extent to which these higher-level patterns may mask meaningful heterogeneity at the level of communities or neighborhoods (13). Similarly, while estimates of all-cause excess mortality have highlighted shifts in overall risks of death during the pandemic, such as racially differential decreases in overall life-expectancy (14), by combining across broad categories of mortality they may obscure increased risks of ARI death that were offset by decreased deaths from other causes, e.g. car accidents, during periods of lockdown or intensified social distancing (15).

## All-cause excess mortality during the COVID-19 pandemic in Michigan

SARS-CoV-2 was first detected in Michigan on March 10, 2020 (16), one day before the World Health Organization declared COVID-19 a global pandemic (17). On the same day, the State of Michigan issued executive orders implementing non-pharmaceutical interventions (NPIs) aimed at limiting in-person interaction to stem the spread of SARS-CoV-2 (18–20). By the end of 2022, > 3 million SARS-CoV-2 cases and > 40,000 COVID-19 deaths had been officially reported in Michigan (21). In the analysis presented here, we used fine-scale spatiotemporal mortality data from Michigan to characterize spatial and temporal variation in excess mortality attributable to acute respiratory infections (ARIs) and to compare this to spatiotemporal shifts in mortality from all other infectious and noncommunicable causes of death (non-ARI deaths).

In Michigan, as well as many other settings, SARS-CoV-2 incidence and mortality rates have been shown to vary along the contours of socioeconomic and geographic inequities which impact healthcare access (22), vaccine uptake (23), employment in higher-risk ‘essential work’ (24), and other risks disproportionately experienced by lower-income populations and people of color. To assess the importance of these drivers, we rely on the widely used CDC/ATSDR Social Vulnerability Index (SVI) (25). Because spatial aggregation of these risk factors may hide important pockets of vulnerability (26), we also examined whether associations between tract-level social vulnerability status and excess mortality (25), differed meaningfully from the same set of associations made at the county level.

In addition, it has been documented that some causes of death, including non-ARI causes such as car crashes (15) and suicides (27) and ARI causes like influenza (28), in fact decreased in Michigan throughout the COVID-19 pandemic. Because of this, all-cause excess mortality estimates that capture the net changes in these broader categories may inadvertently mask the direct burden of SARS-CoV-2 infection-related death, which is of primary interest for surveillance and intervention. To understand the implications of further aggregating these data across spatial locations, we also used estimates obtained at relatively granular level of census tracts as our ‘ground truth’ and then compared results obtained at this level to county-level estimates. This allowed us to quantify the extent to which spatial aggregation can obscure between-location variation and hinder public health policymaking and intervention.

## Methods

For the time interval from December 2019-November 2022, we quantified ARI and non-ARI excess mortality in Michigan at two spatial levels: 1) the relatively coarse level of counties and, 2) the more granular level of census tracts. To evaluate how ARI and non-ARI causes of death varied over time, we divided the SARS-CoV-2 pandemic (12/2019-11/2022) into seven different phases associated with key policy and epidemiological changes (Table 1). For each of these phases, we estimated rates of excess ARI, non-ARI, and all-cause mortality, and computed descriptive statistics indicating the level of spatial aggregation in each of these outcomes.

**Table 1.**
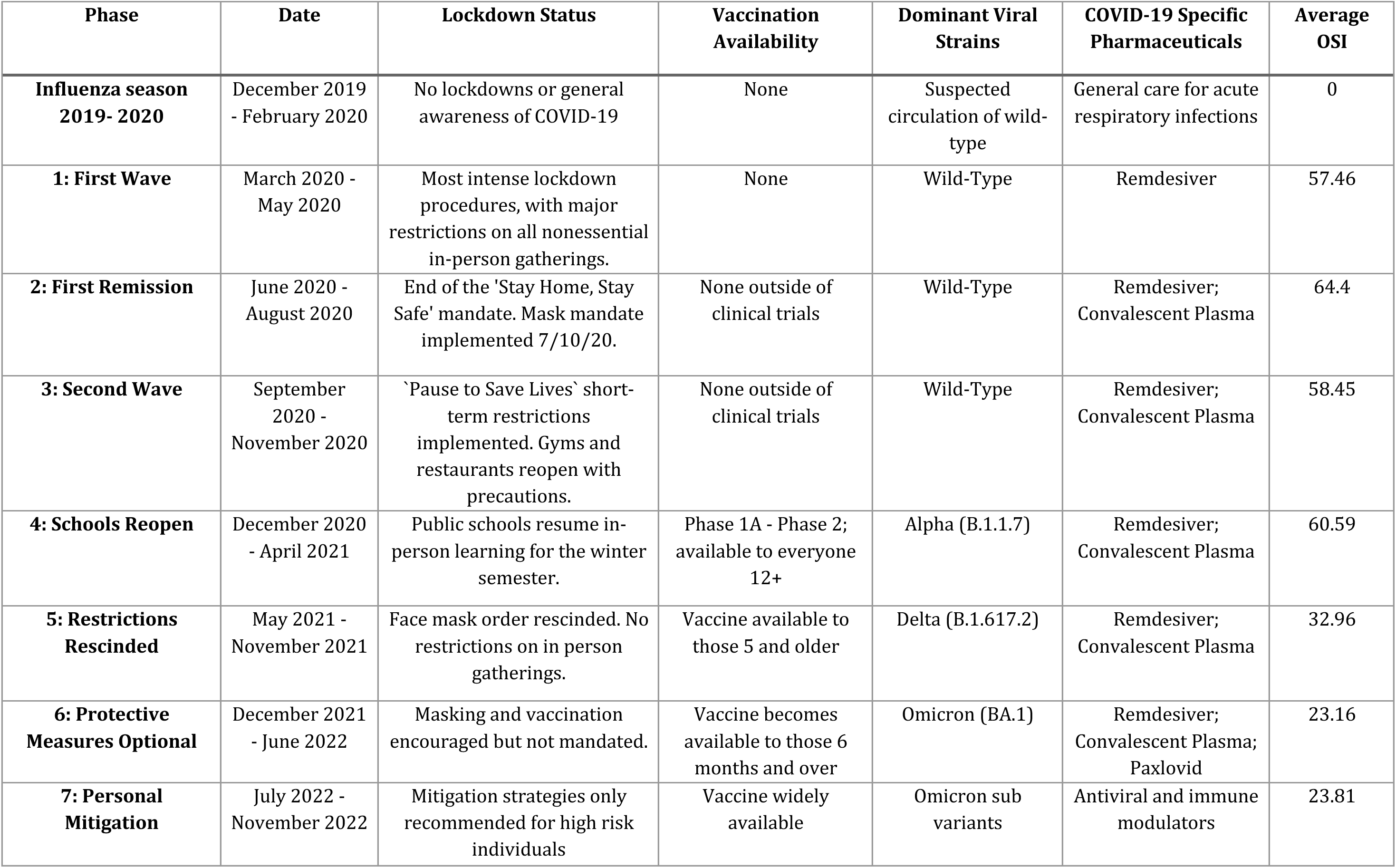
Temporal divisions of the pre-pandemic and pandemic periods, based on key pandemic mitigation strategies.

| Phase | Date | Lockdown Status | Vaccination Availability | Dominant Viral Strains | COVID-19 Specific Pharmaceuticals | Average OSI |
| --- | --- | --- | --- | --- | --- | --- |
| **Influenza season 2019- 2020** | December 2019 - February 2020 | No lockdowns or general awareness of COVID-19 | None | Suspected circulation of wild-type | General care for acute respiratory infections | 0 |
| **1: First Wave** | March 2020 - May 2020 | Most intense lockdown procedures, with major restrictions on all nonessential in-person gatherings. | None | Wild-Type | Remdesiver | 57.46 |
| **2: First Remission** | June 2020 - August 2020 | End of the 'Stay Home, Stay Safe' mandate. Mask mandate implemented 7/10/20. | None outside of clinical trials | Wild-Type | Remdesiver; Convalescent Plasma | 64.4 |
| **3: Second Wave** | September 2020 - November 2020 | 'Pause to Save Lives' short-term restrictions implemented. Gyms and restaurants reopen with precautions. | None outside of clinical trials | Wild-Type | Remdesiver; Convalescent Plasma | 58.45 |
| **4: Schools Reopen** | December 2020 - April 2021 | Public schools resume in-person learning for the winter semester. | Phase 1A - Phase 2; available to everyone 12+ | Alpha (B.1.1.7) | Remdesiver; Convalescent Plasma | 60.59 |
| **5: Restrictions Rescinded** | May 2021 - November 2021 | Face mask order rescinded. No restrictions on in person gatherings. | Vaccine available to those 5 and older | Delta (B.1.617.2) | Remdesiver; Convalescent Plasma | 32.96 |
| **6: Protective Measures Optional** | December 2021 - June 2022 | Masking and vaccination encouraged but not mandated. | Vaccine becomes available to those 6 months and over | Omicron (BA.1) | Remdesiver; Convalescent Plasma; Paxlovid | 23.16 |
| **7: Personal Mitigation** | July 2022 - November 2022 | Mitigation strategies only recommended for high risk individuals | Vaccine widely available | Omicron sub variants | Antiviral and immune modulators | 23.81 |

## Data

### Mortality and sociodemographic data

Individual-level records for all deaths reported in the state of Michigan from January 1, 2015 – November 30, 2022, were obtained from the Vital Records and Health Statistics Division of the Michigan Department of Health and Human Services (MDHHS). Out of 770,831 death records, 93% included complete information on residential address, age, and an ICD-10 coded primary cause of death. Deaths prior to 2017 were originally coded in the ICD-9 scheme and were converted to ICD-10 classification by MDHHS. To focus on mortality risk differences between older and younger individuals, we dichotomized age into two categories: ≤ 65 and 65+. Deaths missing spatial data (52,892 records, details in SI Figure 1) were excluded from this analysis. We divided causes of death into those classified as ARIs and those from all other causes based on ICD-10 cause of death (see SI Table 1 for included ICD-10 codes included in our ARI definition). Annual age-specific population data for each census tract were obtained from the American Community 5-year Survey (ACS-5) from 2015- 2022 (29). Census tract and county level social vulnerability metrics were obtained from the CDC/ATSDR Social Vulnerability Index (SVI) (25). For additional information on deaths included in this analysis see Supplementary Figure 1.

**Figure 1.**
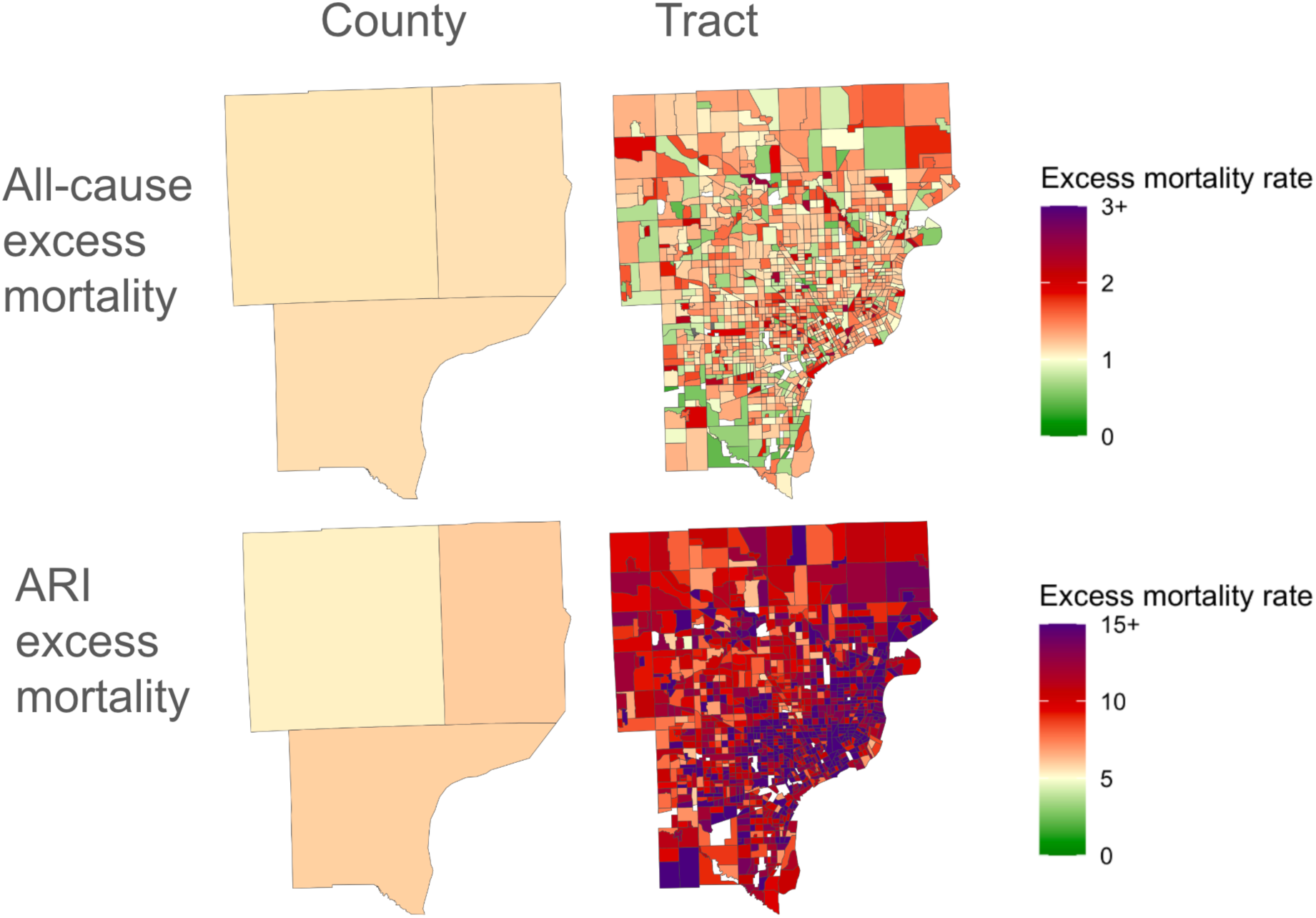
Difference in excess mortality rate estimates across different spatial units and causes of death. The figure displays the impact of aggregation from the census tract to county level on spatial variation in ARI and all-cause excess mortality in the Detroit metropolitan area (Macomb, Oakland, and Wayne counties), from March-May 2020, when ARI-associated excess mortality was at its highest level during the SARS-CoV-2 pandemic.

### Pandemic phases

We divided the time interval from 2/2020-11/2022 into 7 distinct phases of the SARS-CoV-2 pandemic, which we determined using information on the strength of NPIs, as measured by the Oxford Stringency Index (OSI) (30), vaccine availability, COVID-19 specific pharmaceutical treatments (e.g., monoclonal antibodies, antivirals such as Paxlovid and Remdesivir), and shifts in dominant SARS-CoV-2 variants. Table 1 contains detailed information about the specific features of each pandemic phase. We included one pre-pandemic phase from 12/2019–2/2020, covering the majority of the 2019-2020 influenza seasons, to compare pandemic effects to those of seasonal influenza. We used deaths from 1/2015 –11/2019 to estimate seasonal baseline mortality for all models and label it the “reference period” in further analyses

## Statistical Models

Following Beaney (7), we used a two-stage process to estimate excess mortality: First, using records of all deaths in Michigan from 1/2015-11/2019, we estimated the expected number of deaths per calendar month by age group and cause of death category (ARI, non-ARI, and all-cause) for census tract and county divisions of our data (*baseline* mortality). We then compared the observed mortality rate (ARI, non-ARI, all-cause) for the corresponding time period during the SARS-CoV-2 pandemic to our baseline expected mortality to obtain excess mortality estimates in the form of excess mortality rates (EMRs), or the number of observed deaths per the number of expected deaths based on our modeling of historical data.

### Baseline mortality

To capture census tract and county-level variation in pre-pandemic ARI and non-ARI mortality, we used a hierarchical Bayesian negative binomial regression model to model monthly mortality rates by each census tract (Model 1) and county (Model 2; additional information on model selection and fit in Supplementary Information). A random intercept was included to account for unobserved heterogeneity within each of the spatial units (2913 census tracts; 83 counties). Categorical age and year relative to 2015 were included as fixed effects, as shown in Equation 1, below:

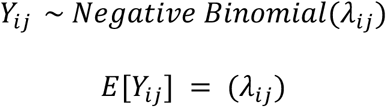

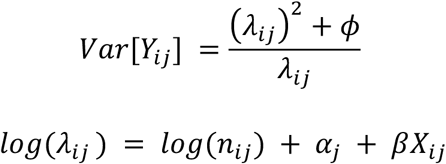

Eq. 1 defines a hierarchical regression model for characterizing variation across census tracts and age groups (dichotomized as 65 and under or 65+). *Y_i_*_j_ represents the number of deaths for the i- th observation of the j-th spatial unit as a function of β, a vector of coefficients representing effects that are stationary across census tracts corresponding to measured covariates, **X**, including the decedent age group (dichotomized as 65≤, 65+), month, and year. The unit-level random intercept, *α*_j_, accounts for unobserved sources of variation in mortality within spatial units. Finally, Φ is an over-dispersion parameter describing how much observed variance exceed the expected variance for a binomial distribution.

The population in units of 100,000 individuals of spatial unit *j* at time *i* is denoted as *n_i_*_j_ and included as an offset term. This allows our covariate estimates to reflect per-100K rates. Using our baseline mortality model fit to data from the pLPre-pandemic period from 1/2015- 11/2019, we predicted expected deaths from ARI and non-ARI causes for each census tract (Model 1) and county (Model 2) by month, year, and age group during the pre-pandemic phase and all pandemic phases from 12/2019-11/2022 as the focus of our analysis. We then aggregated the number of predicted monthly deaths in each census tract or county into the epidemiologically meaningful pandemic periods we have defined above.

### Excess mortality

The excess mortality rate was calculated as the ratio of the observed to expected mortality rate during each pandemic phase, again using a negative binomial regression with the expected number of deaths, derived from Equation 1, included as an offset term. Phase was included as a fixed effect and random effect was included for the spatial unit. This model was employed for each of the six aggregation scenarios of mortality cause and spatial unit. We additionally modeled this relationship with percentile of social vulnerability, as measured by the CDC/ATSDR social vulnerability index so that we could examine the association between a 10 percentile change in social vulnerability variation in excess mortality. To allow the effects of social vulnerability on excess mortality to vary by over time, we included an interaction term between social vulnerability percentile and phase, with the pre-pandemic phase covering the 2019-2020 pre-pandemic influenza season used as a reference. All multilevel regression analyses were conducted in R v.4.2.2 with Bayesian hierarchical modeling conducted using the package *brms* v.2.19 (31,32).

### Spatial variation in excess mortality

We estimated excess mortality rates for ARI, non-ARI, and all-cause excess mortality during each phase at both the census tract and county level in Table 2. We measured spatial aggregation of mortality using the Theil index (*q*), a general entropy metric that summarizes the difference between a uniform spatial distribution of mortality(*q* = 0), versus the maximum aggregation of deaths into a small number of spatial units (*q* = ∞). We employed the Theil index because it is additively decomposable, which allows us to examine the relative contributions of local (census tract) vs. intermediate (county) levels to statewide aggregation (33).

**Table 2.**
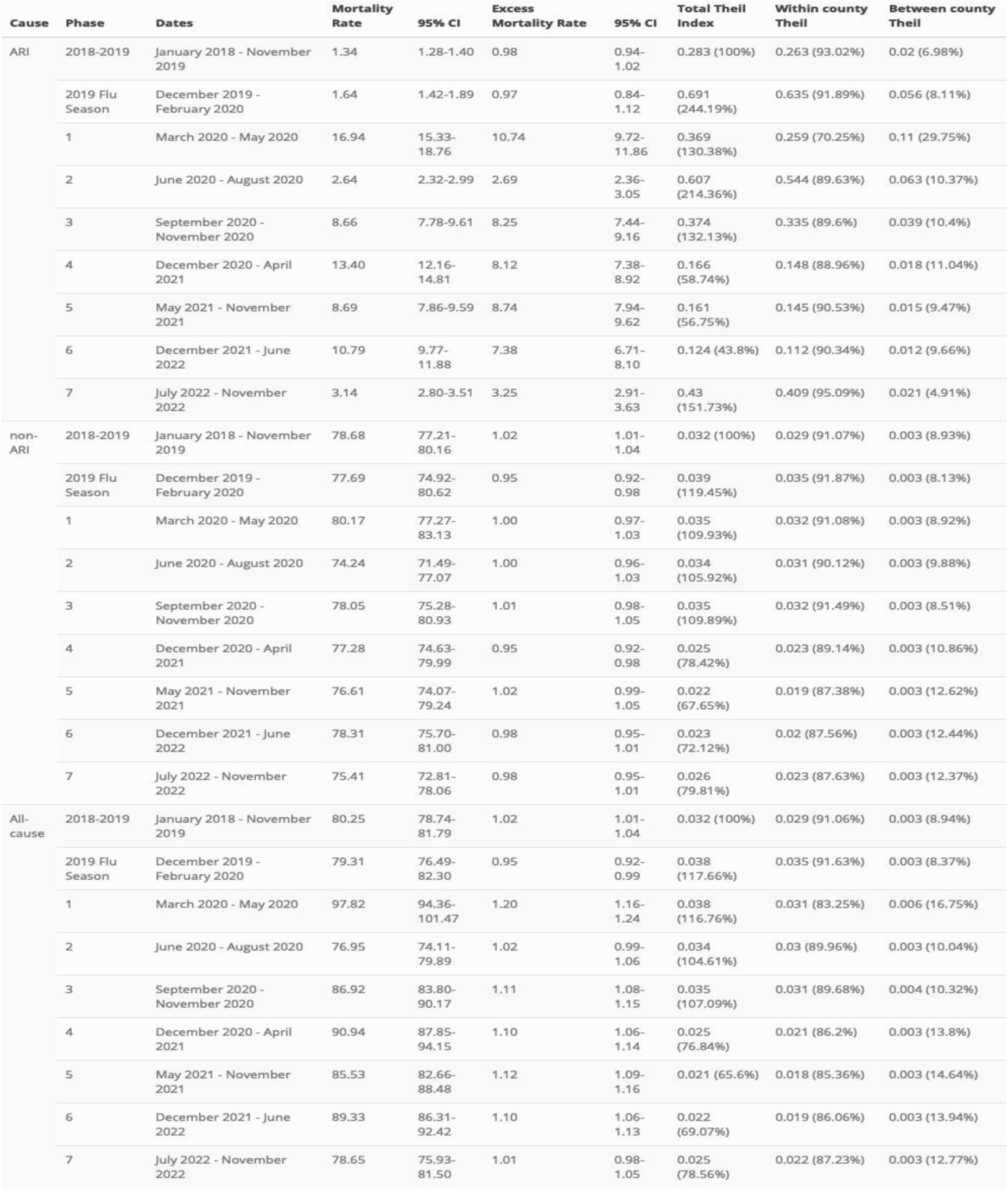
Spatiotemporal variation in ARI, non-ARI, and all-cause mortality and excess mortality. . Average census tract level mortality and excess mortality rates are displayed by phase. Mortality rates are reported as average monthly deaths per 10,000 people. Excess mortality rates are reported as the average monthly census tracts deaths per 1 expected death. The total Theil index is presented, and the percent as compared to phase 1 follows. The decomposed Theil index is presented as overall magnitude and the percent of variation occurring within- versus between-counties during a given pandemic phase.

## Results

From January 2015 to November 2022 there were 813,219 deaths recorded in Michigan, of which 760,327 (93.5%) had geospatial information. During the pre-pandemic reference period from 1/2015-11/2019, there were 446,289 deaths recorded in Michigan with available location data, with 8,225 (1.8%) of these deaths attributable to ARIs. From December 2019 - November 2022, there were an additional 314,038 deaths, with 32,665 (10.4%) from ARIs (Supplementary Information Figure 1).

### ARI, non-ARI and all-cause mortality rates

During the pre-pandemic period (January 2015- November 2019), the median monthly census-tract level non-ARI mortality rate was 78.7/100,000 residents (95% CI: 77.2 - 80.2), and the ARI mortality rate was 1.3 deaths per 100,000 residents (95% CI: 1.3 - 1.4). ARI mortality increased during the December 2019 to February 2020 period to 1.6 deaths per 100,000 residents (95% CI: 1.5 - 1.8), while non-ARI mortality slightly decreased to 77.7 deaths per 100,000 residents (95% CI: 76.0 - 79.4). Reflecting the toll of death exacted by the pandemic, during phases 1-7, the ARI mortality rate remained significantly elevated, peaking in the first phase of the pandemic at 16.9 deaths per 100,000 residents (95% CI: 16.3 - 17.6) and reaching its lowest level in phase 2 at 2.6 (95% CI: 2.5 - 2.8). The non-ARI mortality rate also peaked in phase 2 at 80.2 deaths per 100,000 residents (95% CI: 78.5 - 81.9) but remained below the reference level for phases 3-7.

### Excess mortality estimates

The ARI excess mortality rate spiked during the first pandemic phase (3/2020-5/2020) to a ratio of 10.7 observed to expected deaths (95% CI: 10.3 - 11.1). Although the ARI EMR relative to the pre-pandemic reference period fell during phase 2 (6/2020-8/2020) when lockdown measures were most stringent (EMR = 2.7, 95% CI: 2.5 - 2.9) this trend did not persist: Instead, the ARI EMR increased to 8.3 deaths per expected death (95% CI: 7.9 –8.7) in phase 3 (9/2020-11/2020) and remained > 7 until phase 7 (7/2022-11/2022) when it dropped to 3.3 (95% CI: 3.0 - 3.4). In comparison, the non-ARI excess mortality rate was 1.02 deaths per expected death during the reference period of 1/2018-11/2019 (95% CI: 1.01- 1.04) and was not significantly elevated or reduced during the first pandemic phase (EMR= 1.00, 95% CI: 0.97 - 1.03). In contrast to the large shifts observed in ARI-associated excess mortality, non-ARI excess mortality was not meaningfully elevated above our historical reference period at any phase during the pandemic and fell slightly below it (0.95, 95% CI: 0.92- 0.98) during Phase 4 (12/2020-4/2021).

Examination of these trends in all-cause excess mortality shows how aggregation across causes may obscure dramatic shifts in ARI mortality: The all-cause excess mortality rate was 1.02 deaths per expected death during the reference period (95% CI: 1.00 - 1.04) and dropped during the 2019 flu season (12/2019 –2/2020) to 0.95 (95% CI: 0.93 -0.97). The all-cause EMR peaked during the first pandemic phase at 1.20 (95% CI: 1.17 - 1.22) and remained elevated above the reference period until phase 7, but at a much lower level as compared to the increases in ARI-only excess mortality.1

### Spatial variability in excess mortality

Figure 1 illustrates patterns of excess ARI and non-ARI mortality for three metropolitan counties in Southeast Michigan. This shows how important local-level variation may be washed out by aggregation up to higher level administrative units. State-wide excess ARI and non-ARI mortality is in Supplementary Information Table 2 To quantify this result we computed Theil index values at the census tract and county levels to characterize the degree to which spatial heterogeneity in excess mortality is characterized by differences at a granular level (between census tracts within counties) or at a coarser one (between counties).

The within-county Theil index, which measures the proportion of total statewide variation in EMRs occurring between census tracts within the same county, accounted for 93% of total heterogeneity in ARI mortality during the reference period (1/2018 – 11/2019). The tract-level share of spatial variation in ARI mortality decreased 70% during the first pandemic phase (3/2020-5/2020), indicating a saturation effect in which spatial patterning of ARI mortality became more similar across counties and census tracts as rates rose statewide. This pattern was consistent but attenuated over time, and within-country heterogeneity was depressed below reference levels until phase 7 (7/2022 – 11/2022) when it returned to 95% of the total heterogeneity within the state. Between-tract heterogeneity was also the dominant source of variation in non-ARI excess mortality, at 91% during the reference period. This remained relatively stable, decreasing to 88% by Phase 7. Within-county heterogeneity accounted for 91% of total variation in the reference period’s all-cause mortality and dropped to 83% in the first pandemic phase (3/2020-5/2020) and continued to account for 85-89% of total heterogeneity for the remainder of the pandemic.

Changes in within- vs between-county heterogeneity are reflected in ARI excess mortality rates when comparing county and tract level estimates (Supplementary Information Table 9). Average tract-level ARI excess mortality peaked during Phase 1 (3/2020-5/2020) at 10.7 (95% CI: 10.3 - 11.1) observed deaths for each expected death, while the county level ARI excess mortality rate was 4.7 (95% CI: 4.1 – 5.4). County-level ARI excess mortality ratios were not predictably related to tract level estimates, ranging from 0.42 times the tract estimate in Phase 1 (3/2020-5/2020) to 1.2 times the tract estimate in Phase 5 (5/2021-11/2021). However, county level non-ARI and all-cause excess mortality ratios were consistently within 15% of the tract level excess mortality ratios (Appendix Table 2).

### Census tract characteristics predict excess mortality

Increasing SVI percentile – representing a relative increase in social vulnerability as compared to other census tracts in Michigan - was associated with increasing excess mortality from ARI causes during the reference period (1/2015-11/2019), with an excess mortality rate ratio (EMRR) of 1.04 (95% CrI: 1.03 - 1.06) associated with a 10% increase in proportion non-white. The EMRR associated with a 10% change in SVI percentile increased to 1.1 (95% CrI: 1.08 - 1.13) in the first pandemic phase (3/2020-5/2020), remaining elevated though Phase 3 (IRR: 1.0, 95% CrI: 0.98 -1.03), when lockdowns were most stringent, as measured by the OSI (30). Uncertainty intervals for the EMRR associated with a 10% increase in SVI included the null value of 1 during Phases 6 and 7 (12/2021-11/2022). While spatial aggregation from tract to county did not have a major impact on the estimated relationship between social vulnerability and excess ARI mortality, combining ARI and non-ARI deaths did attenuate the estimated effect of social vulnerability: At the tract level, a 10% increase in SVI was associated with only a relatively small increase in all-cause excess mortality during the first pandemic phase, peaking at an EMRR of 1.02 (95% CrI: 1.02 - 1.03).

## Discussion

Our findings indicate that using county-level all-cause excess mortality as a proxy for ARI excess mortality may obscure the burden of ARI death, and that these effects may be most pronounced during the most acute phases of an emerging epidemic or pandemic. Using data aggregated to a coarse spatial level, such as county, can compound this problem by effectively cancelling out the high ARI mortality in certain tracts by combining them with nearby, low-mortality tracts. Our results suggest that tracts where this high rate of excess ARI mortality is masked by lower rates in nearby areas are likely to have the highest levels of social vulnerability, potentially obscuring the needs of those most at risk.

For example, during the most severe phase of the COVID-19 pandemic (3/2020-5/2020) in Michigan, relying on all-cause mortality to gauge the extent of SARS-CoV-2 deaths would have resulted in a nearly 90% undercount due in part to a corresponding decrease in non-ARI excess mortality during a period of heightened non-pharmaceutical interventions, as calculated using excess mortality rates in Table 1. During the COVID-19 pandemic, large changes in ARI excess mortality were effectively masked by inverse trends in non-ARI excess mortality, as seen in Table 2. While the high infection rate may have indirectly reduced non-infection deaths via increased non-pharmaceutical interventions, this should not be considered a policy success. But combining these negatively correlated outcomes into a single number may result in an incorrectly optimistic assessment of the utility of an ineffective intervention. To monitor future infectious disease outbreaks, surveillance systems should take advantage of the careful categorization of deaths in the hierarchical ICD-10 framework to create metrics that maximize sensitivity and limit the dependence on shifting testing practices. This is an extension of current efforts in syndromic surveillance both in Michigan’s Syndromic Surveillance System and nationally.

Across all phases of the COVID-19 pandemic analyzed here, excess ARI mortality was spatially concentrated, as reflected in Figure 1 and the Theil index calculations in Table 2, which indicate that variability in ARI excess mortality was greater within- than between counties.

Additionally, ARI mortality was more spatially concentrated than the excess mortality from non-ARI causes. This may account for the reduced sensitivity of excess mortality analyses that combine cause-of-death categories and use aggregated spatial categories such as counties. Excess mortality from ARI saw much larger between-county variation in phase 1 than any other phase. This could be due differential rates of introduction of SARS-CoV-2 into the more densely populated areas of Southeast Michigan, prior to consistent statewide spread.

Despite the high burden of mortality exacted by SARS-CoV-2, non-ARI causes still accounted for most of the deaths in our dataset during the COVID-19 pandemic. Because of this, all-cause excess mortality estimates were consistently closer to non-ARI excess mortality than to those of ARI excess mortality. At the tract level, the ARI excess mortality rate was nearly an order of magnitude – 9 times - larger than the all-cause excess mortality rate during the first phase of the pandemic. Detailed data on illness and mortality at the level of individual census tracts, like those we have presented here, are not always available. However, we found that using county-level data alone introduced unpredictable bias in ARI excess mortality rate estimates, from 0.42 times the phase 1 tract EMR to 1.32 times the phase 3 EMR. From left to right, Figure 1 demonstrates the differences in excess mortality estimates in phase 1 when using different spatial units.

Our results also suggest that tract-level clustering of SARS-CoV-2 deaths persisted well into the later phases of the pandemic, when SARS-CoV-2 had become a less urgent concern. For example, Phase 7 (7/2022-11/2022), which most closely represents the current lack of mitigation measures, saw an increase in total within-county aggregation of excess mortality. As shown in Table 2, non-ARI excess mortality became less spatially concentrated, suggesting that the burden of non-ARI excess mortality remains more evenly distributed across the state while the burden of ARI excess mortality has become more concentrated within groups of census tracts again. While it is not clear what explains these patterns, they may be attributable to trends such as the increased prevalence and spatial aggregation of vaccine hesitancy in Michigan (23) and ongoing inequity in access to SARS-CoV-2 mitigation during high-risk periods.

The social vulnerability index of the tract or county was strongly predictive of ARI and non-ARI excess mortality during Phases 1 and 2 (3/2020-8/2020). Following Phase 2, SVI became less predictive of excess mortality, suggesting that the first wave of COVID-19 disproportionately affected socially marginalized communities, as shown in Table 2 and echoing results of previous analyses such as those by Luck et al. (34,35). The effect was much stronger on ARI excess mortality compared to non-ARI excess mortality, demonstrating the negative impacts of COVID-19 were more strongly associated with social vulnerability than the effects of mitigation strategies. Many of these relationships become more attenuated or even inverted over time. These shifts in the relationship between social vulnerability and excess mortality may be attributable to differential adherence to personal mitigation strategies such as work from home, avoidance of high-risk locations, and vaccination. Future studies should investigate the role of specific components of the social vulnerability index, such as race and income, to better understand the underlying causes of excess mortality.

When using the 5-year history of ARI and non-ARI mortality at the census tract level to estimate and characterize fine-scale variability in pre-pandemic expected mortality, we observed how these disaggregated metrics contribute to the combined measure of all-cause excess mortality. Relying on this aggregated metric of all-cause mortality clearly limits the power to detect large shifts in cause-specific mortality, particularly when they follow different trends, and that limitation can be mitigated by even using coarse categories such as ARI vs. non-ARI deaths. Spatial aggregation, while often convenient or made necessary by data privacy restrictions, further limits the sensitivity of surveillance systems and masks important social patterns (36).

The standard practice of aggregating analyses to state or county levels can blind public health practitioners to a concentration of vulnerability that would otherwise be easily visible through qualitative examination of the type of mortality data presented in Figure 1.

From a policy perspective, ARI excess mortality is a convenient target metric for pan-respiratory infectious disease surveillance: It is more sensitive to shifts in ARI-related death than all-cause mortality, but is not dependent on widespread active surveillance or laboratory confirmation of infection, while providing potentially actionable insights about local burden when calculated at the census tract level. Additionally, this division of ARI and non-ARI related deaths can measure which pandemic countermeasures result in changes in non-ARI deaths. In our analysis, we found no evidence of excess non-ARI deaths but aggregation across non-ARI causes could mask individual trends similar to the aggregation of ARI and non-ARI mortality.

There are logistical challenges to working with this type of detailed data and the required collaborations can take time to develop. In technical terms we expect spatially detailed mortality analyses based on grouped ICD-10 to become a common part of epidemiological practice.

## Data Availability

Data can be requested from the Michigan Department of Health and Human Services

## Supplementary Information

**SI Figure 1.**
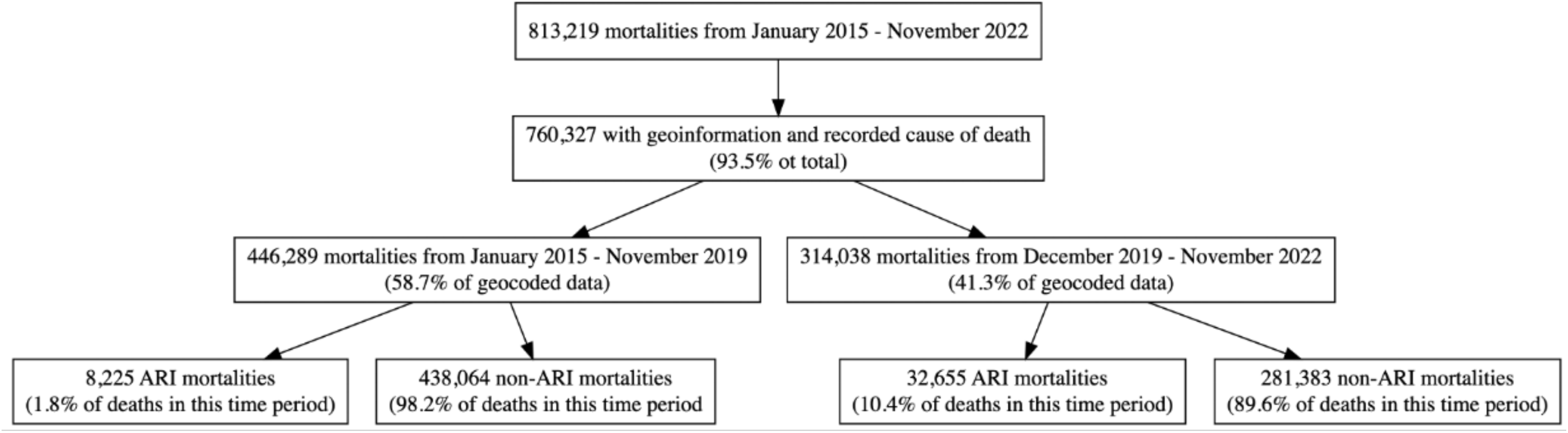
Pipeline of data included in the analysis. Deaths without geoinformation were excluded from the analysis (6.5% of all deaths from January 2015 - November 2022).

**SI Figure 2.**
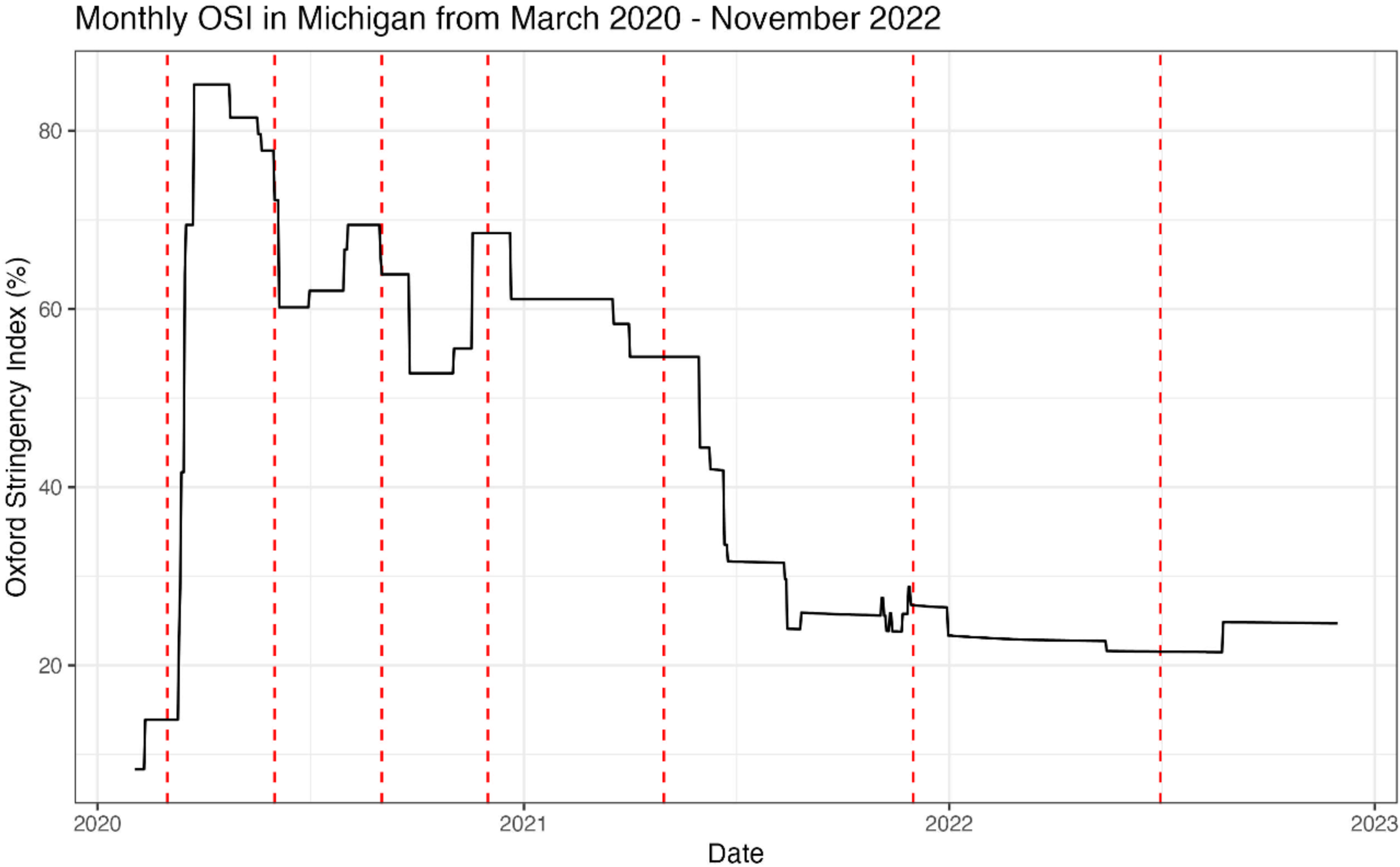
The Oxford stringency index (OSI) is a measure of pandemic mitigation intensity, with higher values indicating greater mitigation, divided amongst the pandemic periods.

**SI Figure 3.**
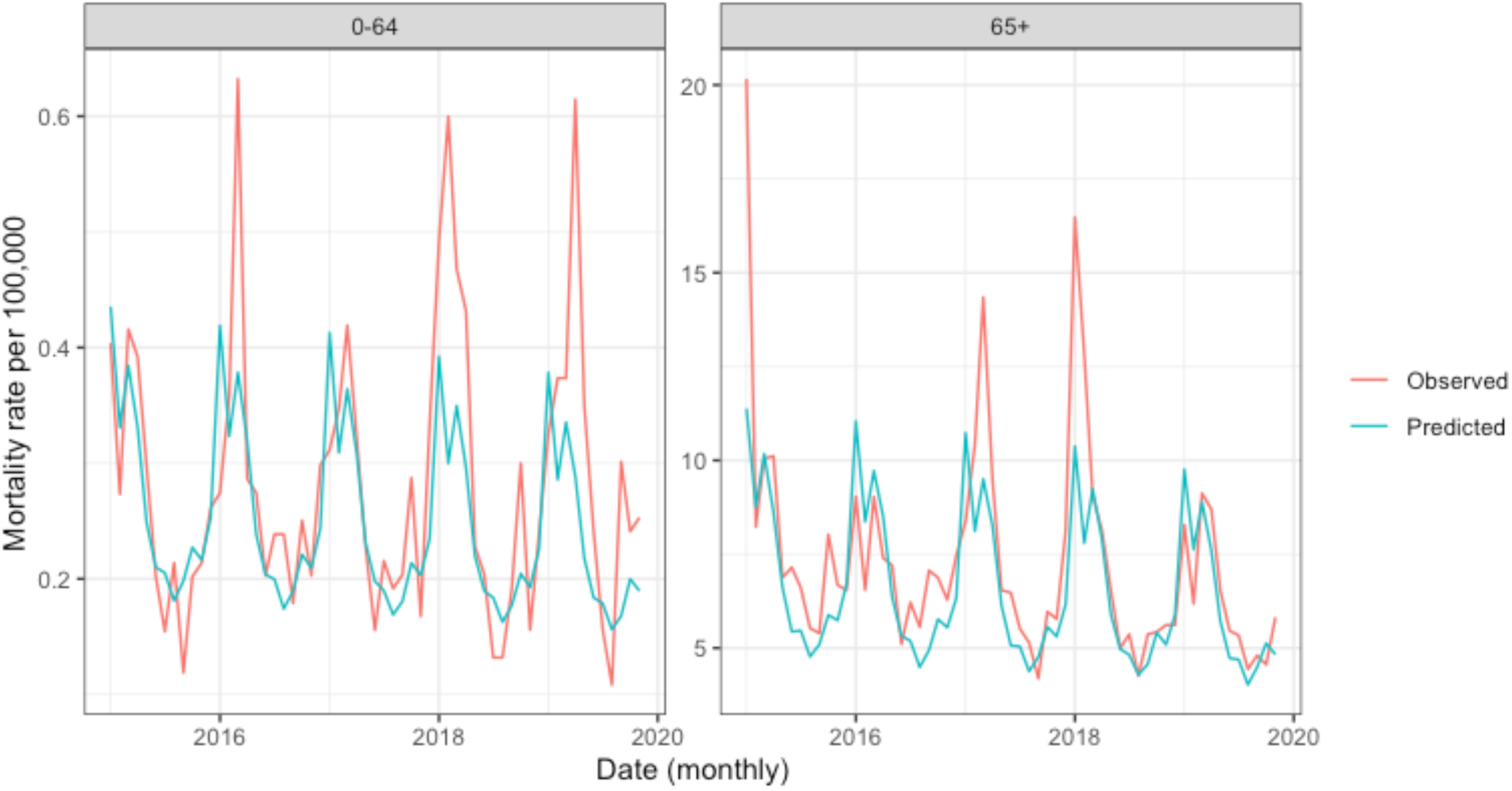
Statewide aggregations of tract level predicted and observation mortality per 100,000 people during the baseline period for ARI causes of death.

**SI Figure 4.**
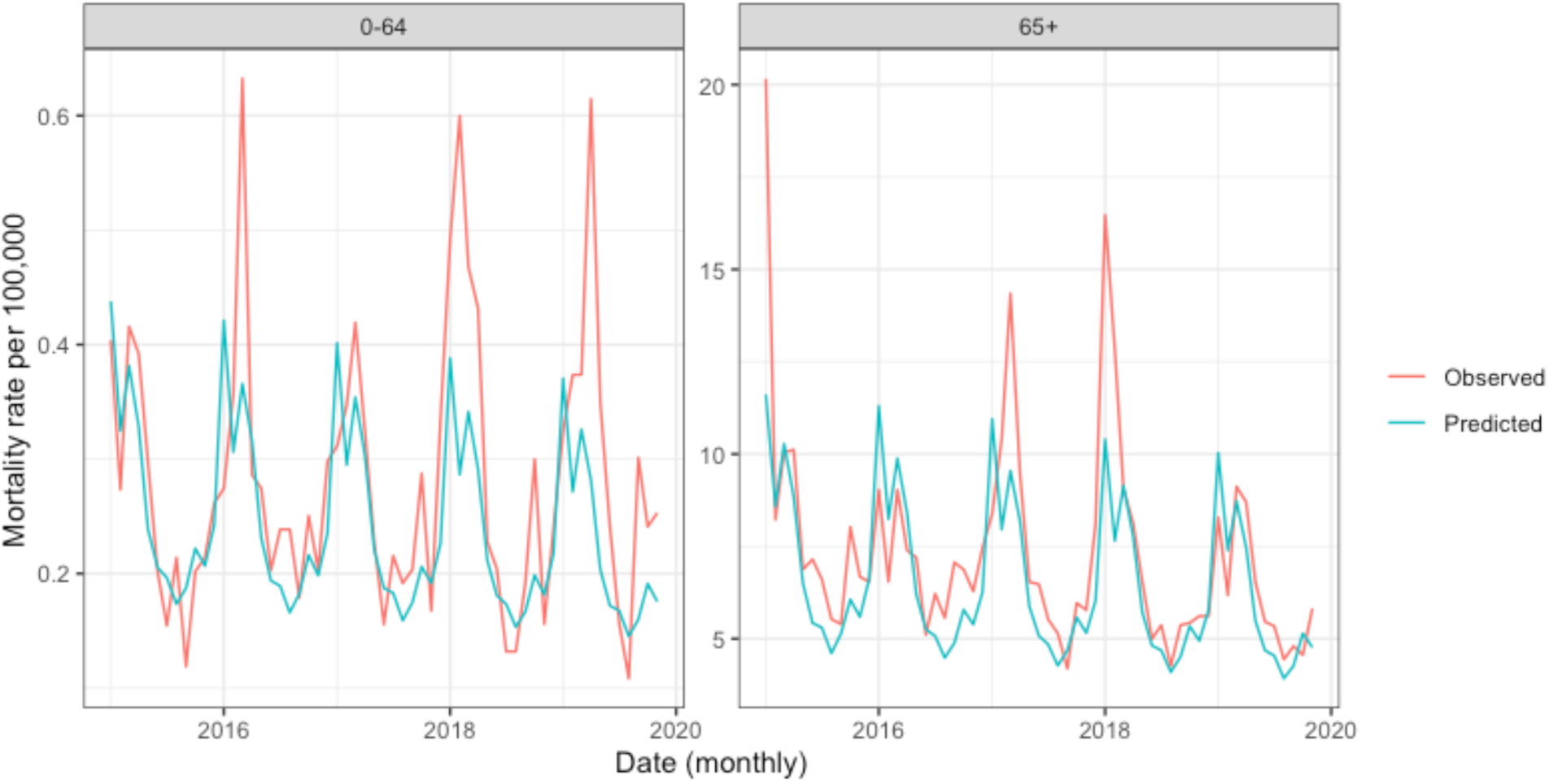
Statewide aggregations of county level predicted and observation mortality per 100,000 people during the baseline period for ARI causes of death.

**SI Figure 5.** Statewide aggregations of tract level predicted and observation mortality per 100,000 people during the baseline period for non-ARI causes of death.

**SI Figure 6.**
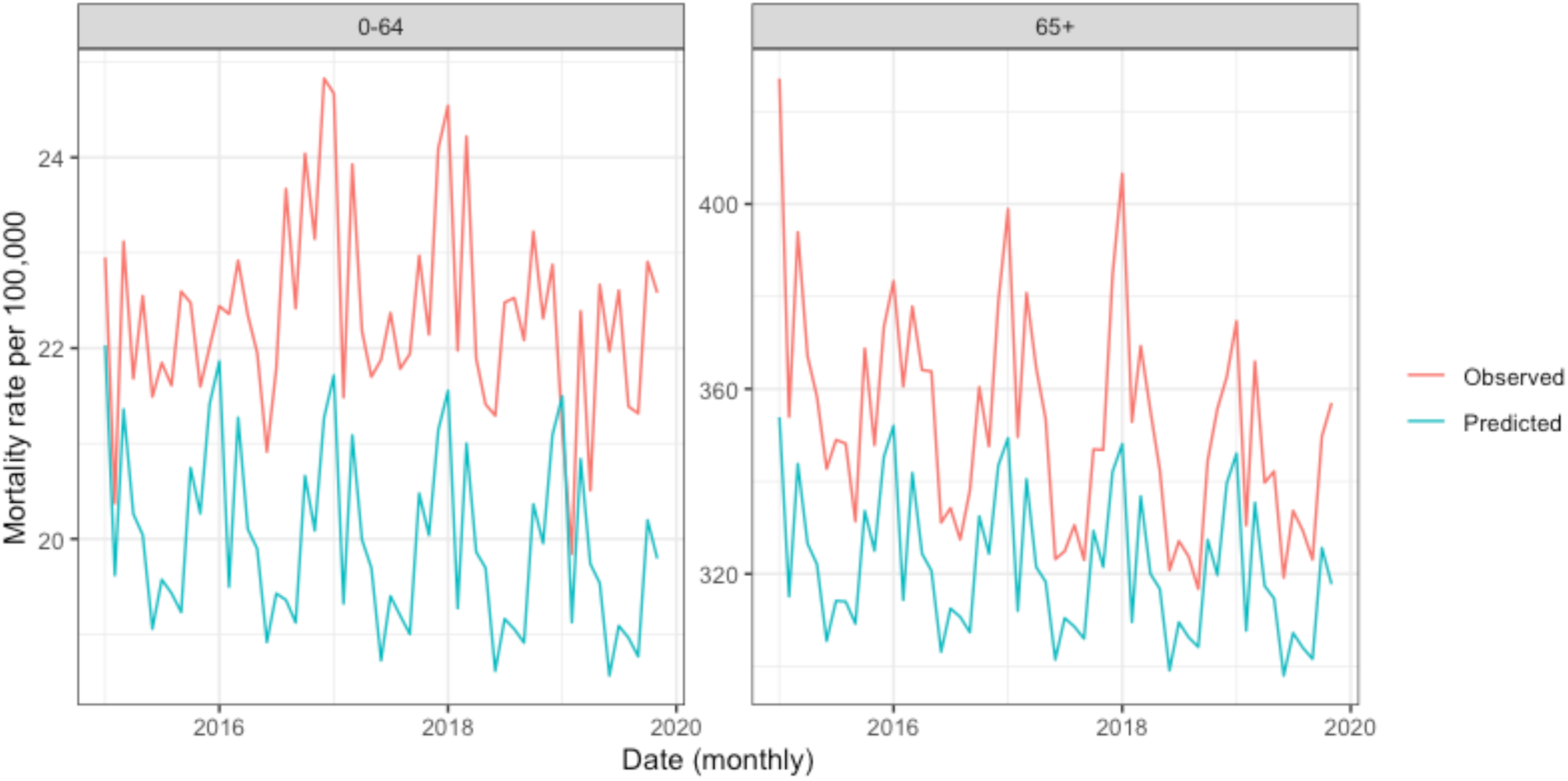
Statewide aggregations of predicted and observation mortality per 100,000 people during the baseline period for non-ARI causes of death.

**SI Figure 7.**
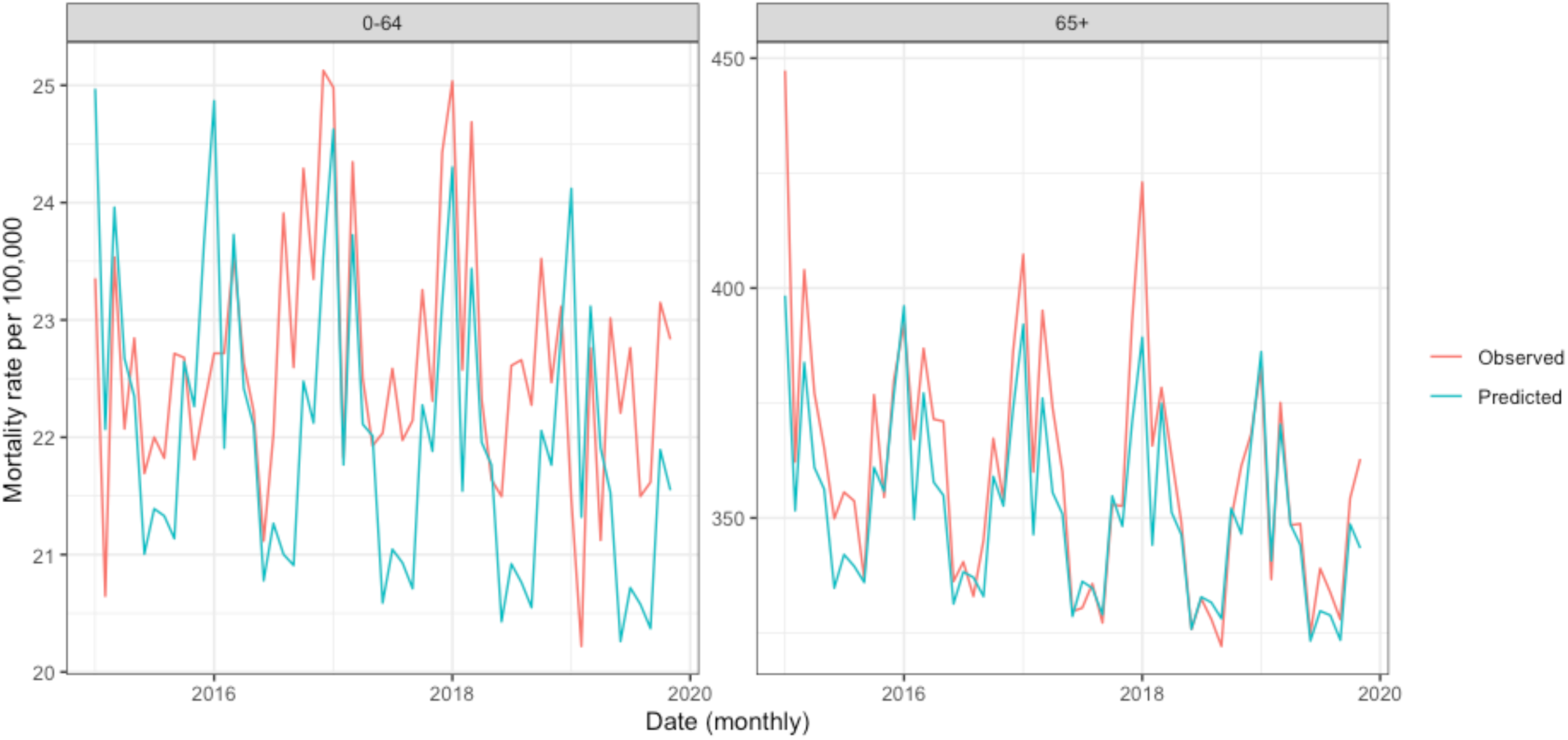
Statewide aggregations of tract level predicted and observation mortality per 100,000 people during the baseline period for all causes of death.

**SI Figure 8.**
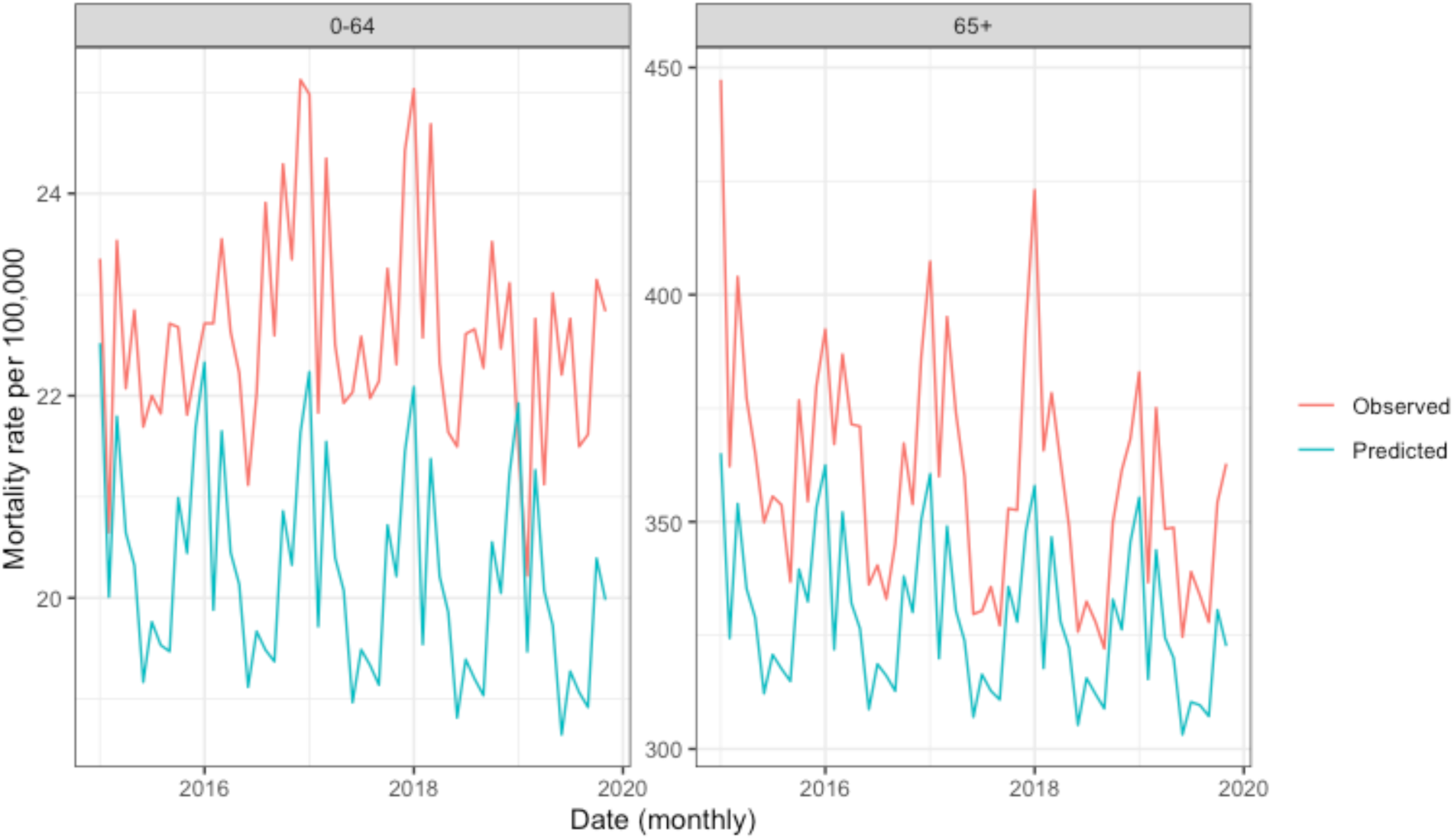
Statewide aggregations of country level predicted and observation mortality per 100,000 people during the baseline period for all causes of death.

**SI Table 1.** ICD-10 codes classified as acute respiratory infections (ARI). All other ICD-10 codes are classified as non-ARI.

| **ICD-10**<br>**Code** | **Description** |
| --- | --- |
| **J010** | Acute maxillary sinusitis |
| **J020** | Streptococcal pharyngitis |
| **J029** | Acute pharyngitis, unspecified |
| **J050** | Acute obstructive laryngitis [croup] |
| **J051** | Acute epiglottitis |
| **J069** | Acute upper respiratory infection, unspecified |
| **J09** | Influenza due to certain identified influenza viruses |
| **J100** | Influenza due to other identified influenza virus with pneumonia |
| **J101** | Influenza due to other identified influenza virus with other respiratory manifestations |
| **J108** | Influenza due to other identified influenza virus with other manifestations |
| **J110** | Influenza due to unidentified influenza virus with pneumonia |
| **J111** | Influenza due to unidentified influenza virus with other respiratory manifestations |
| **J118** | Influenza due to unidentified influenza virus with other manifestations |
| **J120** | Adenoviral pneumonia |
| **J121** | Respiratory syncytial virus pneumonia |
| **J122** | Parainfluenza virus pneumonia |
| **J123** | Human metapneumovirus pneumonia |
| **J128** | Other viral pneumonia |
| **J129** | Viral pneumonia, unspecified |
| **J13** | Pneumonia due to *Streptococcus pneumoniae* |
| **J14** | Pneumonia due to *Hemophilus influenzae* |
| **J150** | Pneumonia due to *Klebsiella pneumoniae* |
| **J151** | Pneumonia due to *Pseudomonas* |
| **J152** | Pneumonia due to staphylococcus |
| **J153** | Pneumonia due to streptococcus, group B |
| **J154** | Pneumonia due to other streptococci |
| **J155** | Pneumonia due to *Escherichia coli* |
| **J156** | Pneumonia due to other Gram-negative bacteria |
| **J157** | Pneumonia due to *Mycoplasma pneumoniae* |
| **J158** | Pneumonia due to other specified bacteria |
| **J159** | Unspecified bacterial pneumonia |
| **J180** | Bronchopneumonia, unspecified organism |
| **J181** | Lobar pneumonia, unspecified organism |
| **J182** | Hypostatic pneumonia, unspecified organism |
| **J188** | Other pneumonia, unspecified organism |
| **J189** | Pneumonia, unspecified organism |
| **J206** | Acute bronchitis due to rhinovirus |
| **J208** | Acute bronchitis due to other specified organisms |
| **J209** | Acute bronchitis, unspecified |
| **J210** | Acute bronchiolitis due to respiratory syncytial virus |
| **J218** | Acute bronchiolitis due to other specified organisms |
| **J219** | Acute bronchiolitis, unspecified |
| **J22** | Unspecified acute lower respiratory infection |
| **J80** | Acute respiratory distress syndrome |
| **J81** | Pulmonary edema |
| **J82** | Pulmonary eosinophilia, not elsewhere classified |
| **J840** | Alveolar and parieto-alveolar conditions |
| **J841** | Other interstitial pulmonary diseases with fibrosis |
| **J848** | Other specified interstitial pulmonary diseases |
| **J849** | Interstitial pulmonary disease, unspecified |
| **J850** | Gangrene and necrosis of lung |
| **J851** | Abscess of lung with pneumonia |
| **J852** | Abscess of lung without pneumonia |
| **J860** | Pyothorax with fistula |
| **J869** | Pyothorax without fistula |
| **J90** | Pleural effusion, not elsewhere classified |
| **J930** | Spontaneous tension pneumothorax |
| **J931** | Other spontaneous pneumothorax |
| **J938** | Other pneumothorax and air leak |
| **J939** | Pneumothorax, unspecified |
| **J941** | Fibrothorax |
| **J942** | Hemothorax |
| **J948** | Other specified pleural conditions |
| **J949** | Pleural condition, unspecified |
| **J980** | Diseases of bronchus, not elsewhere classified |
| **J981** | Pulmonary collapse |
| **J982** | Interstitial emphysema |
| **J983** | Compensatory emphysema |
| **J984** | Other disorders of lung |
| **J985** | Diseases of mediastinum, not elsewhere classified |
| **J986** | Disorders of diaphragm |
| **J988** | Other specified respiratory disorders |
| **J989** | Respiratory disorder, unspecified |
| **U071** | COVID-19 |
| **U049** | COVID-19 |

**SI Table 2.** Excess mortality ratios by phase for different spatial and causal categories, and the ratio of county estimates to tract estimates for each causal category.

| Cause | Phase | Tract Estimate | Tract 95% CI | County Estimate | County 95% CI | County/Tract |
| --- | --- | --- | --- | --- | --- | --- |
| ARI | 2018-2019 Reference Period | 0.98 | 0.94-1.02 | 0.96 | 0.87-1.07 | 0.98 |
|  | 2019 Flu Season | 0.99 | 0.89-1.09 | 0.98 | 0.82-1.17 | 0.99 |
|  | Phase 1 | 10.95 | 10.37-11.58 | 4.69 | 4.07-5.4 | 0.43 |
|  | Phase 2 | 2.74 | 2.52-2.98 | 2.46 | 2.09-2.89 | 0.90 |
|  | Phase 3 | 8.42 | 7.93-8.94 | 11.13 | 9.67-12.81 | 1.32 |
|  | Phase 4 | 8.28 | 7.87-8.71 | 9.32 | 8.15-10.63 | 1.13 |
|  | Phase 5 | 8.92 | 8.47-9.39 | 10.85 | 9.5-12.38 | 1.22 |
|  | Phase 6 | 7.53 | 7.16-7.91 | 7.94 | 6.94-9.04 | 1.05 |
|  | Phase 7 | 3.31 | 3.1-3.55 | 3.63 | 3.13-4.23 | 1.10 |
| Non-ARI | 2018-2019 Reference Period | 1.02 | 1.01-1.04 | 1.00 | 0.99-1.02 | 0.98 |
|  | 2019 Flu Season | 0.93 | 0.91-0.95 | 1.01 | 0.99-1.04 | 1.09 |
|  | Phase 1 | 1.17 | 1.15-1.19 | 1.12 | 1.1-1.15 | 0.96 |
|  | Phase 2 | 1.00 | 0.98-1.02 | 1.08 | 1.05-1.11 | 1.08 |
|  | Phase 3 | 1.09 | 1.07-1.11 | 1.22 | 1.18-1.25 | 1.12 |
|  | Phase 4 | 1.07 | 1.06-1.09 | 1.18 | 1.15-1.21 | 1.10 |
|  | Phase 5 | 1.10 | 1.08-1.11 | 1.20 | 1.17-1.23 | 1.09 |
|  | Phase 6 | 1.07 | 1.05-1.09 | 1.16 | 1.14-1.19 | 1.08 |
|  | Phase 7 | 0.99 | 0.97-1.01 | 1.08 | 1.06-1.11 | 1.09 |
| All-cause | 2018-2019 Reference Period | 1.02 | 1.01-1.04 | 1.00 | 0.99-1.01 | 0.98 |
|  | 2019 Flu Season | 0.93 | 0.92-0.95 | 1.01 | 0.99-1.03 | 1.09 |
|  | Phase 1 | 0.98 | 0.96-1 | 1.05 | 1.02-1.07 | 1.07 |
|  | Phase 2 | 0.98 | 0.96-0.99 | 1.06 | 1.04-1.08 | 1.08 |
|  | Phase 3 | 0.99 | 0.97-1.01 | 1.09 | 1.07-1.11 | 1.10 |
|  | Phase 4 | 0.93 | 0.92-0.95 | 1.02 | 1-1.04 | 1.10 |
|  | Phase 5 | 0.99 | 0.98-1.01 | 1.08 | 1.06-1.09 | 1.09 |
|  | Phase 6 | 0.96 | 0.94-0.97 | 1.04 | 1.02-1.06 | 1.08 |
|  | Phase 7 | 0.96 | 0.95-0.98 | 1.04 | 1.03-1.06 | 1.08 |

**SI Table 3.** The effect estimates of a 10% change in social vulnerability index and excess mortality by phase of the pandemic, spatial unit, and cause of death.

| Spatial unit | Phase | ARI |  | non-ARI |  | All-Cause |  |
| --- | --- | --- | --- | --- | --- | --- | --- |
|  |  | IRR | 95% CI | IRR | 95% CI | IRR | 95% CI |
| Tract | Reference | 1.04 | 1.02-1.06 | 1.00 | 1.00-1.01 | 1.00 | 1.00-1.01 |
| Tract | 2019 Flu Season | 1.05 | 1.01-1.08 | 1.00 | 1.00-1.01 | 1.00 | 0.99-1.01 |
| Tract | Phase 1 | 1.10 | 1.08-1.13 | 1.02 | 1.02-1.03 | 1.01 | 1.00-1.01 |
| Tract | Phase 2 | 1.06 | 1.03-1.09 | 1.01 | 1.00-1.01 | 1.00 | 1.00-1.01 |
| Tract | Phase 3 | 1.01 | 0.98-1.03 | 1.00 | 0.99-1.01 | 1.00 | 0.99-1.00 |
| Tract | Phase 4 | 1.03 | 1.01-1.05 | 1.01 | 1.00-1.01 | 1.00 | 1.00-1.01 |
| Tract | Phase 5 | 1.03 | 1.01-1.05 | 1.01 | 1.00-1.01 | 1.00 | 1.00-1.01 |
| Tract | Phase 6 | 1.02 | 1.00-1.03 | 1.01 | 1.00-1.01 | 1.01 | 1.00-1.01 |
| Tract | Phase 7 | 0.99 | 0.97-1.01 | 1.00 | 1.00-1.01 | 1.00 | 1.00-1.01 |
| County | Reference | 1.00 | 0.99-1.00 | 1.02 | 0.98-1.05 | 1.00 | 0.99-1.00 |
| County | 2019 Flu Season | 1.00 | 1.00-1.01 | 1.00 | 0.94-1.06 | 1.00 | 0.99-1.01 |
| County | Phase 1 | 1.00 | 1.00-1.01 | 1.03 | 0.98-1.08 | 1.01 | 1.00-1.01 |
| County | Phase 2 | 1.00 | 0.99-1.01 | 1.06 | 1.00-1.12 | 1.00 | 0.99-1.01 |
| County | Phase 3 | 1.00 | 0.99-1.00 | 0.99 | 0.94-1.04 | 1.00 | 0.99-1.01 |
| County | Phase 4 | 1.00 | 1.00-1.01 | 1.00 | 0.96-1.05 | 1.00 | 1.00-1.01 |
| County | Phase 5 | 1.00 | 0.99-1.01 | 1.01 | 0.97-1.06 | 1.00 | 0.99-1.01 |
| County | Phase 6 | 1.01 | 1.00-1.01 | 0.99 | 0.95-1.04 | 1.00 | 1.00-1.01 |
| County | Phase 7 | 1.00 | 1.00-1.01 | 1.01 | 0.96-1.06 | 1.00 | 0.99-1.01 |

